# rECHOmmend: an ECG-based machine-learning approach for identifying patients at high-risk of undiagnosed structural heart disease detectable by echocardiography

**DOI:** 10.1101/2021.10.06.21264669

**Authors:** Alvaro E. Ulloa-Cerna, Linyuan Jing, John M. Pfeifer, Sushravya Raghunath, Jeffrey A. Ruhl, Daniel B. Rocha, Joseph B. Leader, Noah Zimmerman, Greg Lee, Steven R. Steinhubl, Christopher W. Good, Christopher M. Haggerty, Brandon K. Fornwalt, Ruijun Chen

**Author notes:** Corresponding author: Ruijun Chen, MD, 100 North Academy Ave, Danville, PA, 17822-4400.

## Abstract

**Background:** Early diagnosis of structural heart disease improves patient outcomes, yet many remain underdiagnosed. While population screening with echocardiography is impractical, electrocardiogram (ECG)-based prediction models can help target high-risk patients. We developed a novel ECG-based machine learning approach to predict multiple structural heart conditions, hypothesizing that a composite model would yield higher prevalence and positive predictive values (PPVs) to facilitate meaningful recommendations for echocardiography.

**Methods:** Using 2,232,130 ECGs linked to electronic health records and echocardiography reports from 484,765 adults between 1984-2021, we trained machine learning models to predict the presence of any of seven echocardiography-confirmed diseases within one year. This composite label included: moderate or severe valvular disease (aortic/mitral stenosis or regurgitation, tricuspid regurgitation), reduced ejection fraction <50%, or interventricular septal thickness >15mm. We tested various combinations of input features (demographics, labs, structured ECG data, ECG traces) and evaluated model performance using 5-fold cross-validation, multi-site validation trained on one clinical site and tested on 11 other independent sites, and simulated retrospective deployment trained on pre-2010 data and deployed in 2010.

**Findings:** Our composite “rECHOmmend” model using age, sex and ECG traces had an area under the receiver operating characteristic curve (AUROC) of 0.91 and a PPV of 42% at 90% sensitivity at a prevalence of 17.9% for our composite label. Individual disease models had AUROCs ranging from 0.86-0.93 and lower PPVs from 1%-31%. The AUROC for models using different input features ranged from 0.80-0.93, increasing with additional features. Multi-site validation showed similar results to the cross-validation, with an aggregate AUROC of 0.91 across our independent test set of 11 clinical sites after training on a separate site. Our simulated retrospective deployment showed that for ECGs acquired in patients without pre-existing known structural heart disease in a single year, 2010, 11% were classified as high-risk, of which 41% developed true, echocardiography-confirmed disease within one year.

**Interpretation:** An ECG-based machine learning model using a composite endpoint can predict previously undiagnosed, clinically significant structural heart disease while outperforming single disease models and improving practical utility with higher PPVs. This approach can facilitate targeted screening with echocardiography to improve under-diagnosis of structural heart disease.

## Introduction

Patients with structural heart disease carry a high burden of morbidity and mortality, for whom echocardiography holds important evidence-based implications for diagnosis, prognosis, and management.^1–5^ Echocardiography is the primary diagnostic test for many structural conditions, including valvular disease, left ventricular (LV) dysfunction, and various cardiomyopathies.^6–8^ Early diagnosis of structural heart disease improves patient outcomes, yet despite growing indications and availability of echocardiography, these conditions continue to be underdiagnosed.^9–12^ Studies have shown that millions of patients have unrecognized disease, including the majority of elderly patients found to have moderate or severe valvular disease on community screening and the majority of patients with hypertrophic cardiomyopathy.^11,12^

Electrocardiogram (ECG)-based machine learning models can help identify undiagnosed patients for targeted screening, yet limitations to their practical adoption remain. ECGs are more common, inexpensive, and broadly indicated than echocardiograms, and machine learning approaches using ECGs have been shown to identify patients at increased risk of individual diseases.^13–15^ However, despite otherwise good performance, these models often suffer from low positive predictive values (PPVs) due to the low prevalence of individual target diseases.^15,16^ This limits the practical utility of real-world implementations, since many patients identified as high-risk would need to undergo screening to diagnose one true case.

We therefore sought to combine multiple models into a single platform to increase diagnostic yield. We developed a novel machine learning approach to identify patients at high-risk for any of seven structural heart disease endpoints within a single ECG platform, including moderate or severe valvular disease (aortic stenosis [AS], aortic regurgitation [AR], mitral stenosis [MS], mitral regurgitation [MR], tricuspid regurgitation [TR]), reduced left ventricular ejection fraction (EF), and increased interventricular septal (IVS) thickness. Our model generates a composite prediction with higher yield/PPV to facilitate a practical clinical recommendation for diagnostic echocardiography. Moreover, we simulated the utility of this model on a large retrospective dataset to assess expected real-world performance if implemented into clinical care.

## Methods

### Data

The Institutional Review Board approved this study with a waiver of consent. We retrieved and processed data from three clinical sources at Geisinger, a large regional US health system providing both inpatient and outpatient care, including 2,110,332 patients from the Epic (Epic Systems, Madison, WI) electronic health record (EHR), 758,269 echocardiograms from Xcelera (Philips, Cambridge, MA), and 3,548,543 ECGs from MUSE (GE Healthcare). We included all 12-lead ECGs after 1984 from patients ≥ 18 years old, sampled at either 250hz or 500hz, and a corresponding Epic medical record, resulting in 2,925,925 ECGs from 631,710 patients. All data were collected through July 2021.

We obtained vitals, laboratory results, and patient demographics as of the index ECG acquisition date and time (Supplemental Table 1). We used the closest past measurement unless the measurement was older than one year, in which case we assigned a missing value. We extracted echocardiographic measurements and diagnoses from Xcelera reports and ECG structured findings, measurements, and 12-lead traces from MUSE.^13,17^ Structured ECG findings were directly obtained from the final, official interpretation by an attending cardiologist. We then labeled ECGs as detailed below. Overall, we included 2,232,130 ECGs with at least 1 label from 484,765 patients (Figure 1).

**Figure 1:**
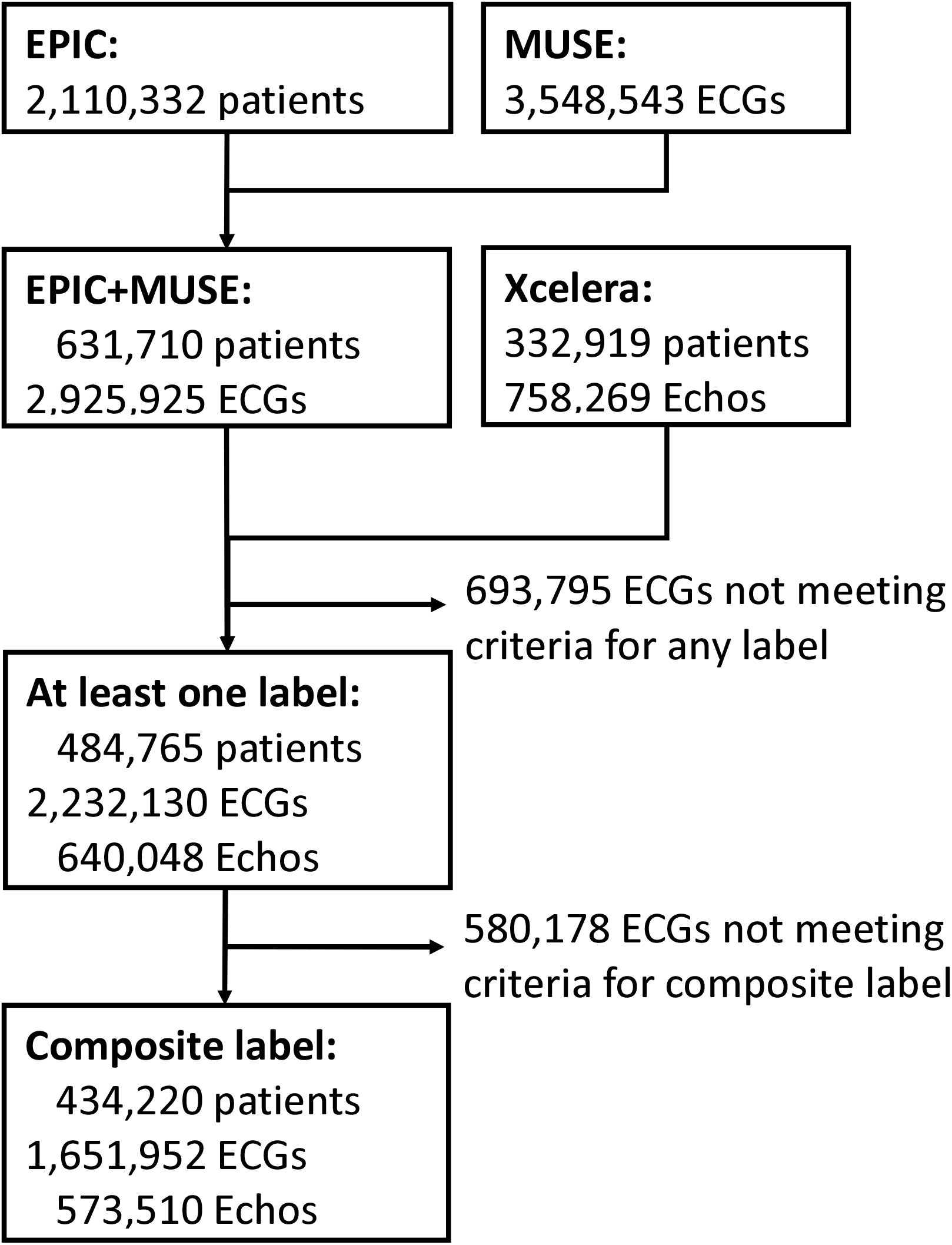
Flow diagram from source data to the dataset used for all experiments. We processed data from research repositories created from electronic health record data from Epic, ECG data from MUSE, and echocardiography data from Xcelera. The clinical MUSE database was processed to include 12-lead ECGs sampled at either 250hz or 500hz, acquired after 1984 from patients older than 18 years of age.

### Echocardiography-confirmed disease outcome definitions

We defined 7 outcome labels using echocardiography reports, one for each disease outcome (AS, AR, MR, MS, TR, reduced EF, increased IVS thickness). We used regular expressions to extract key words and phrases identifying the diagnosis of valvular stenosis or regurgitation and its associated severity level, based upon the final interpretation by an attending cardiologist (Supplemental Table 2). We labeled each of the valvular conditions of interest as *positive* if moderate or severe and *negative* if normal or mild in severity. We assigned a missing label otherwise.

We defined *positive* labels for reduced EF as a reported EF of <50% on echocardiography. We defined increased IVS thickness as >15mm. These criteria were chosen based on cardiologist and clinician consensus and in concordance with existing guidelines for potential diseases of interest, such as hypertrophic cardiomyopathy.^18^ Echocardiograms not meeting those criteria were labeled as *negative*. We assigned a missing label if the measurement was missing.

Outcome labels extracted from echocardiography reports for AS, AR, MR, MS, and TR were randomly sampled in sets of 100-200 and validated by manual chart review.

### ECG labeling

For each given disease outcome, an ECG was labeled as *positive* if it was acquired within one year before the patient’s first positive echocardiography report for that disease, or any time after the echocardiogram until a censoring event (Supplemental Figure 1). Censoring events were defined as death, end of observation in the EHR, or any intervention that directly treated the disease and could modify the underlying physiology, such as valve replacement or repair. We also used a negative echocardiography report after a positive echocardiography report as a censoring event to conservatively eliminate the possibility that such interventions may have been performed at outside institutions and therefore not represented in our data.

For each given disease outcome, an ECG could be labeled as *negative* using 2 sets of criteria, depending on whether the patient did or did not have a history of prior echocardiography. 1) For patients with a prior history of echocardiography, ECGs acquired more than one year prior to the last negative echocardiogram, with confirmed absence of that given disease, were labeled as *negative* (Supplemental Figure 1). 2) In the absence of any patient history of echocardiography, an ECG was also labeled as *negative* if there was at least 1 year of subsequent follow-up without a censoring event and without any coded diagnoses for the relevant disease (Supplemental Table 3).

For the composite endpoint, we labeled an ECG as positive if any of the seven individual outcomes were positive and as negative if all seven outcomes were negative.

### Model Development

We developed 9 models using different combinations of input feature sets from structured data (demographics, vitals, labs, structured ECG findings and measurements) and ECG voltage traces. For ECG trace models, we developed a low-parameter convolutional neural network (CNN) with 18,495 trainable parameters that consisted of six one-dimensional CNN-Batch Normalization-ReLU layer blocks.^19^ The blocks were followed by a two-layer multilayer perceptron and a final logistic output layer (Supplemental Table 4). Each CNN layer consisted of 16 kernels of size 5. We used the same configuration to train one model per clinical outcome, resulting in 7 independently trained CNN models (Figure 2).

**Figure 2:**
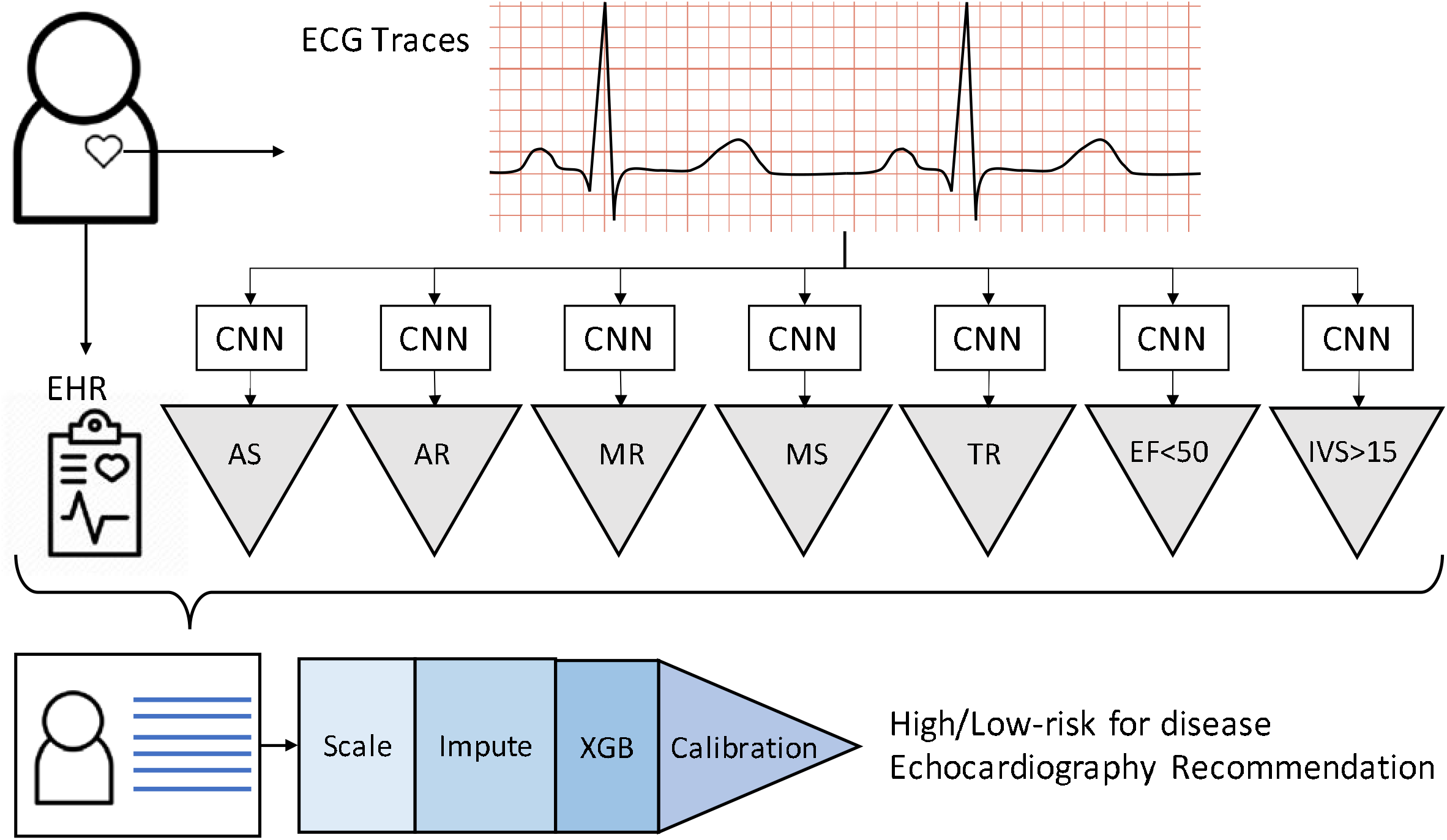
rECHOmmend model diagram showing the classification pipeline for ECG traces and other EHR data. The output (gray triangle) of each convolutional neural network (CNN) applied to ECG trace data is concatenated with labs, vitals, and demographics to form a feature vector. The vector is the input to the classification pipeline (min-max scaling, mean imputation, XGBoost classifier, and calibration), which outputs a composite prediction for the patient.

To form the final model and combine ECG trace-based models with structured data, we concatenated the risk scores from the individual CNNs with the structured data. We used the concatenated feature vector to train a classification pipeline consisting of a min-max scaler (min 0, max 1), mean imputation, XGBoost classifier, and calibration (Figure 2).^20,21^

### Model Evaluation

We evaluated the models using three approaches: 1) a traditional random cross-validation partition; 2) a multi-site validation where the model was trained on data from Geisinger Medical Center and tested on 11 other independent clinical sites; and 3) a retrospective deployment scenario where, using 2010 as the simulated deployment year, we used past data to train and future data to test. We measured AUROC, area under the precision-recall curve (AUPRC), and other performance metrics (sensitivity, specificity, positive and negative predictive values) at multiple operating points. For all experiments, data were split into training, internal validation, and test sets with no overlap of patients across these sets.

### Cross validation

We conducted a 5-fold cross validation by randomly sampling 5 mutually exclusive sets of patients. We expanded each set to all ECGs from each patient to form the training and test sets. When training the CNN models for each individual endpoint, we discarded samples with missing labels. We applied the model to all test samples and evaluated performance only on samples with complete labels that also satisfied the rECHOmmend labeling criteria, described above. Performance statistics were reported as means and 95% confidence intervals (CIs) across five folds for a random ECG per patient.

### Multi-site validation

To perform multi-site validation, we created 12 mutually exclusive sets of patients from the 12 clinical sites in the Geisinger Health System. We assigned each patient to a particular site by selecting the most common ECG site of origin for that given patient. We removed any ECGs taken outside of the assigned site for each patient.

We trained our model on data from patients at a single site—Geisinger Medical Center, a large quaternary teaching hospital in Danville, Pennsylvania. We then tested this model on 11 other independent clinical sites, ranging from outpatient centers to small community hospitals to large teaching hospitals, at various locations across Pennsylvania.

### Retrospective deployment

We retrospectively simulated a deployment of our model using a cutoff date of January 1, 2010, re-labeling all ECGs with information available as of that date. We used this artificially constrained dataset to replicate the cross-validation experiments and train a deployment model using data prior to 2010. We then applied the deployment model to the first ECG per patient for all patients seen from January 1, 2010 through December 31, 2010. We calibrated the XGBoost model using earliest ECGs from the at-risk population in 2005 and measured performance statistics on all patients at risk in 2010. We determined the true outcomes of the at-risk population using information up to July 23, 2021, following the definitions for positive and negative outcome labels outlined above.

### Sensitivity Analyses

To account for potential variation in what providers and patients may find to be clinically significant disease, we repeated the cross-validation experiment on a different set of labels representing severe disease only. These label definitions include severe valvular disease only (moderate valvular disease now considered a negative label) and changed the definition for reduced EF to be <35%.

To account for the possibility of patients with persistently undiagnosed disease in our definition of negative ECGs, we also repeated our cross-validation experiment using only echocardiography-confirmed negatives. Patients who never received an echocardiogram were excluded. All ECGs labeled ‘negative’ were followed by a negative echocardiogram confirming the absence of that given disease outcome.

## Results

We identified 758,269 echocardiography reports from 332,919 patients, of which 191,652 echocardiograms from 88,093 patients were positive for at least one disease outcome label. Disease prevalence ranged from 0.6% for MS to 17.2% for reduced EF (Supplemental Table 5). We identified 2,232,130 ECGs from 484,765 patients who met criteria for at least one positive or negative individual disease label, of which 1,651,952 ECGs from 434,220 patients qualified for the composite label (Supplemental Table 6). At baseline, across 2.23 million ECGs, the median patient age was 64 years, 50.1% were male, and 97.1% were white (Table 1). ECGs from patients with a positive label as compared to a negative label were generally older with a higher proportion of males and smokers. Baseline characteristics among patients with missing or undefined labels as compared to patients with at least one defined label were largely similar (Supplemental Table 7).

**Table 1:** Baseline characteristics and features at time of index ECG, reported as mean (SD) and median [IQR], for continuous values, or percentage, for categorical values. BP, blood pressure. BBB, bundle branch block. COPD, chronic obstructive pulmonary disease. MI, myocardial infarction. PVC, premature ventricular contractions. SVT, supraventricular tachycardia.

|  | Mean<br>(SD) / % | Median [IQR] |  | Mean<br>(SD) / % | Median [IQR] |
| --- | --- | --- | --- | --- | --- |
| **Demographics and Vitals** |  |  | **ECG Findings and Measurements** |  |  |
| Age (years) | 63 (17) | 64 [52, 76] | R Axis | 22 (50) | 21 [-10, 54] |
| BMI (kg/m2) | 31 (9) | 30 [25, 35] | PR Interval | 165 (212) | 160 [144, 182] |
| Systolic BP (mmHg) | 129 (20) | 128 [116, 140] | P Axis | 48 (30) | 50 [33, 64] |
| Diastolic BP (mmHg) | 73 (12) | 72 [64, 80] | QRS Duration | 98 (25) | 90 [82, 104] |
| Heart Rate (bpm) | 76 (15) | 74 [66, 84] | QT | 400 (51) | 398 [368, 430] |
| Height (cm) | 168 (11) | 168 [160, 178] | QTC | 445 (54) | 440 [418,464] |
| Weight (kg) | 88 (24) | 85 [70, 101] | T Axis | 52 (53) | 46 [23, 71] |
| Race (%White) | 97.1% |  | Ventricular Rate | 77 (20) | 74 [63, 87] |
| Sex (%Male) | 50.1% |  | Avg RR Interval | 821 (194) | 814 [688, 946] |
| Smoker (%Ever) | 59.7% |  | Normal | 43.8% |  |
| **Labs** |  |  | Prior Infarct | 18.7% |  |
| A1C (%) | 6.9 (3) | 6.5 [5.8, 7.5] | Non-Specific T-wave Changes | 16.0% |  |
| Bilirubin (mg/dL) | 0.57 (0.60) | 0.5 [0.3,0.7] | Sinus Bradycardia | 14.1% |  |
| BUN (mg/dL) | 20.5 (12.8) | 17 [13, 23] | Non-Specific ST Changes | 10.3% |  |
| Cholesterol (mg/dL) | 172 (47) | 168 [140, 200] | Ischemia | 10.0% |  |
| CKMB (ng/mL) | 8.9 (32.2) | 2.9 [1.9,5] | Left Axis Deviation | 9.3% |  |
| Creatinine (mg/dL) | 1.2 (1.4) | 0.9 [0.8,1.2] | Atrial Fibrillation | 8.5% |  |
| CRP (mg/L) | 36.2 (63.9) | 9 [2.6,38] | Left Ventricular Hypertrophy | 8.0% |  |
| D dimer (mcg/mL) | 1.5 (2.6) | 0.6 [0.3, 1.5] | Tachycardia | 7.5% |  |
| Glucose (mg/dL) | 119 (48) | 104 [93, 125] | Prior Anterior MI | 7.3% |  |
| HDL (mg/dL) | 48 (16) | 45 [37, 56] | PVC | 6.8% |  |
| Hemoglobin (g/dL) | 14 (34) | 13 [11.7, 14.3] | First Deg Block | 6.3% |  |
| LDH (U/L) | 249 (237) | 207 [171, 264] | Right BBB | 6.0% |  |
| LDL (mg/dL) | 95 (38) | 91 [68,117] | Prolonged QT | 5.0% |  |
| Lymphocytes (%) | 23 (11) | 22 [15, 29] | Poor Tracing | 4.9% |  |
| Potassium (mmol/L) | 4.2 (0.7) | 4.2 [3.9, 4.5] | Premature Atrial Contractions | 4.8% |  |
| Pro-BNP (pg/mL) | 5002 (10668) | 1369[341,4377] | Pacemaker | 4.6% |  |
| Sodium (mmol/L) | 139 (3) | 140[137, 141] | T-wave Inversion | 4.6% |  |
| Troponin I (ng/mL) | 1 (13) | 0.03 [0.01,0.06] | Low QRS voltage | 4.4% |  |
| Troponin T (ng/mL) | 0.16 (0.84) | 0.01 [0.01,0.04] | Fascicular Block | 3.2% |  |
| Triglyceride (mg/dL) | 154 (122) | 127 [90, 183] | Incomplete Right BBB | 3.1% |  |
| Uric Acid (mg/dL) | 6.6 (2.4) | 6.3 [4.9,7.9] | Left BBB | 2.8% |  |
| VLDL (mg/dL) | 29 (16) | 25 [18, 36] | Intraventricular Block | 2.3% |  |
| eGFR (mL/min/1.73m2) | 54 (12) | 60 [55, 60] | Right Axis Deviation | 2.2% |  |
| **Other Comorbidities** |  |  | Atrial Flutter | 1.3% |  |
| Heart Failure | 17.2% |  | Acute MI | 1.0% |  |
| Prior MI | 18.8% |  | Incomplete left BBB | 0.4% |  |
| Diabetes Mellitus | 23.1% |  | Supraventricular Tachycardia | 0.4% |  |
| COPD | 14.0% |  | Early Repolarization | 0.3% |  |
| Renal Failure | 8.3% |  | Complete Heart Block | 0.1% |  |
| Prior Echocardiogram | 28.4% |  | Other Bradycardia | 0.1% |  |
| Coronary Artery Disease | 23.1% |  | Second-degree AV block | 0.1% |  |
| Hypertension | 46.4% |  | Ventricular Tachycardia | 0.1% |  |

### Model Input Feature Evaluation

Table 2 shows the results of 5-fold cross validation comparing model performance as a function of different input features. AUROCs ranged from 0.80 for the model using only age and sex to 0.93 for the model with all available inputs, including structured ECG findings and measurements, demographics, labs, vitals, and ECG traces (Figure 3). While the model with all available inputs provided the best performance, we focus the remainder of our results in this manuscript on models that include only age, sex, and ECG traces since this input set best balances portability, objectivity, and performance, with an AUROC of 0.91. These inputs are all directly available from MUSE or other ECG systems, without additional integration with other data sources, and do not require waiting for the official cardiologist interpretation, which may be subject to inter-rater variability. Complete, detailed results including all other input sets for every disease label across all folds and various subgroups are available at: http://rechommend.herokuapp.com/.

**Table 2:** Performance comparison of cross-validated models across various input features for the composite endpoint (valvular disease, reduced EF, increased IVS). All values are shown in percentages with the 95% CI in between brackets. Each model was tested based on a random ECG per patient. AUROC, area under receiver operating curve. AUPRC, area under precision-recall curve. PPV, positive predictive value. ECG, electrocardiogram. EHR, electronic health record.

| Input Features | AUROC | AUPRC | PPV (%) @ 90% Sensitivity | Specificity (%) @ 90% Sensitivity |
| --- | --- | --- | --- | --- |
| A) Age + Sex | 0.799 [0.795,0.802] | 0.468 [0.462,0.473] | 27.5 [27.0,28.0] | 48.2 [47.5,49.0] |
| B) Demographics, Labs, and Vitals | 0.862 [0.860,0.865] | 0.651 [0.644, 0.657] | 32.3 [31.8,32.8] | 58.9 [58.3,59.5] |
| C) ECG Structured Findings and Measurements | 0.879 [0.877,0.881] | 0.677 [0.672, 0.683] | 34.0 [33.4,34.5] | 61.8 [61.0,62.6] |
| D) ECG Traces | 0.904 [0.902,0.906] | 0.719 [0.714, 0.724] | 41.1 [40.4,41.9] | 71.9 [71.3,72.6] |
| **Available from ECG system** |  |  |  |  |
| Age + Sex + ECG Traces | 0.907 [0.905, 0.908] | 0.714 [0.707, 0.722] | 42.0 [41.4,42.6] | 72.9 [72.4,73.4] |
| C + D | 0.912 [0.910, 0.913] | 0.739 [0.733, 0.744] | 42.9 [42.0,43.8] | 73.9 [73.2,74.6] |
| **Available from ECG + EHR** |  |  |  |  |
| A + B + C | 0.917 [0.915, 0.919] | 0.762 [0.757, 0.767] | 44.2 [43.5,44.9] | 75.2 [74.6,75.8] |
| A + B + D | 0.925 [0.923, 0.926] | 0.780 [0.775, 0.784] | 46.7 [46.0,47.4] | 77.6 [77.0,78.2] |
| A + B + C + D | 0.928 [0.927, 0.930] | 0.787 [0.783, 0.792] | 47.8 [47.2,48.4] | 78.6 [78.2,79.0] |

**Figure 3.**
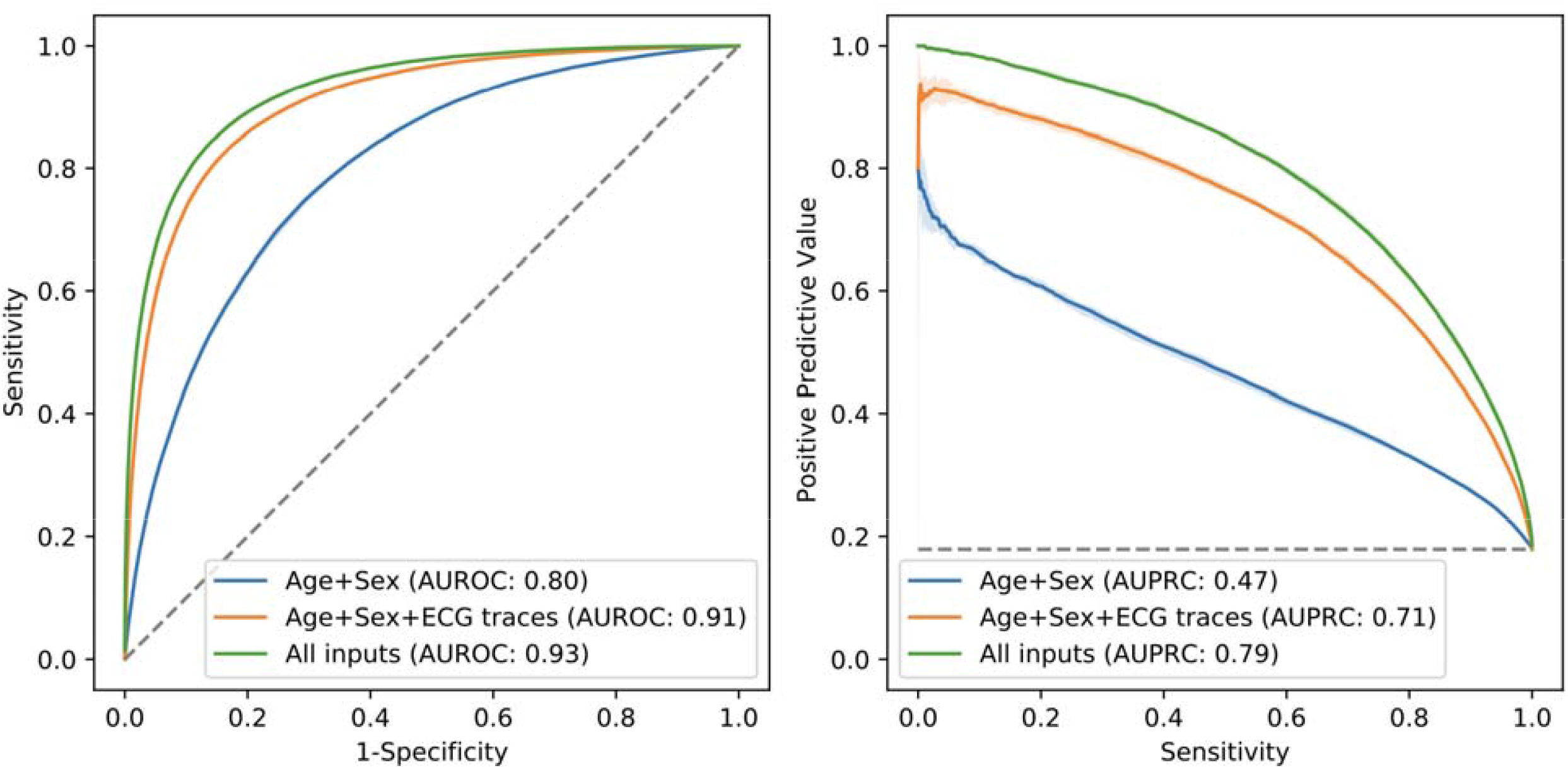
Performance of the rECHOmmend model in cross-validation experiments across various inputs. The figure on the left shows the area under the receiver operating curve (AUROC) while the figure on the right shows the area under the precision-recall curve (AUPRC).

### Cross-validation performance of rECHOmmend model

The rECHOmmend model using age, sex, and ECG traces for prediction of the composite disease label yielded an AUROC of 0.91 [95% CI 0.90, 0.91] and a PPV of 42% at 90% sensitivity with 18% disease prevalence (Table 3). As hypothesized, the composite model yielded a higher PPV than any of the 7 models trained for an individual component endpoint, which ranged from 1% for MS to 31% for reduced EF (Table 3). We found the same trend for the AUPRC, 0.71 [95% CI 0.71, 0.72] for the rECHOmmend model, as compared to individual model AUPRCs, which ranged from 0.04-0.65 (Supplemental Figure 2). Performance metrics for alternate model operating points are presented in Supplemental Table 8.

**Table 3:** Age + Sex + ECG traces model results for cross-validation experiments for each individual disease outcome and composite rECHOmmend model. Results are shown based on a random ECG per patient and averaged across 5 folds. All values are shown in percentages with the 95% CI in between brackets. AUROC, area under receiver operating curve. AUPRC, area under precision-recall curve. PPV, positive predictive value.

| **Disease** | **Prevalence (%)** | **AUROC** | **AUPRC** | **PPV (%) @ 90% Sensitivity** | **Specificity (%) @ 90% Sensitivity** |
| --- | --- | --- | --- | --- | --- |
| Aortic Stenosis | 2.4 [2.3,2.5] | 0.908 [0.900, 0.915] | 0.221 [0.204, 0.239] | 8.4 [7.7,9.1] | 75.7 [73.6,77.7] |
| Aortic Regurgitation | 1.8 [1.8,1.9] | 0.849 [0.844, 0.855] | 0.120 [0.114, 0.127] | 3.9 [3.6,4.2] | 58.9 [57.2,60.7] |
| Mitral Regurgitation | 4.5 [4.4,4.6] | 0.911 [0.908, 0.914] | 0.367 [0.347, 0.388] | 15.2 [14.7,15.7] | 76.4 [75.8,77.0] |
| Mitral Stenosis | 0.3 [0.2,0.3] | 0.918 [0.905, 0.930] | 0.039 [0.036, 0.044] | 1.1 [1.0,1.3] | 79.4 [75.3,82.9] |
| Tricuspid Regurgitation | 4.7 [4.6,4.9] | 0.915 [0.909, 0.920] | 0.415 [0.393, 0.438] | 16.1 [14.7,17.7] | 76.9 [74.7,78.9] |
| EF<50% | 9.2 [9.1,9.2] | 0.929 [0.926, 0.931] | 0.647 [0.633, 0.662] | 31.4 [30.2,32.7] | 80.2 [79.1,81.2] |
| IVS>15mm | 4.0 [3.9,4.1] | 0.862 [0.856, 0.868] | 0.223 [0.213, 0.234] | 9.4 [8.8,10.1] | 64.2 [61.7,66.6] |
| rECHOmmend (composite) | 17.9 [17.8,18.0] | 0.907 [0.905, 0.908] | 0.714 [0.707, 0.722] | 42.0 [41.4,42.6] | 72.9 [72.4,73.4] |

### Multi-site validation performance

The rECHOmmend model trained on Geisinger Medical Center and validated across 11 other clinical sites performed similarly well to our cross-validation experiment, yielding an AUROC of 0.91 in aggregate across all other sites (Supplemental Table 9). Individual site AUROCs ranged from 0.79 at the Viewmont Imaging Center to 0.93 at the Scranton Community Medical Center, with 9 out of 11 sites having AUROCs > 0.85 and 8 out of 11 sites having AUROCs ≥ 0.90. The prevalence of the composite label for disease among sites varied from 1% at Viewmont to 39% at the Geisinger Commonwealth School of Medicine (GCSM). Correspondingly, PPV varied from 15% at Viewmont to 54% at GCSM.

### Simulated deployment performance

We identified 692,273 ECGs with a qualifying label for any of the seven clinical outcomes prior to 2010, of which 485,469 ECGs qualified for the composite label to train the deployment model. A cross-validation experiment for this pre-2010 subset showed similar, yet slightly reduced performance as compared with the full dataset (AUROC 0.89; PPV 31% at 90% sensitivity; Supplemental Table 10).

The 2010 deployment test set contained ECGs from 69,544 patients (Figure 4A). After excluding patients with a known history of disease, we identified 63,459 at-risk patients between January 1 and December 31, 2010. Of these patients, outcome labels for 20,395 were undefined due to inadequate follow-up or not meeting criteria for the composite label. As previously noted, the characteristics of patients with undefined labels were similar to those with defined labels. The AUROC among patients with defined labels was 0.86. Using a threshold estimated to yield 90% sensitivity based on the pre-2010 training data, the deployment model labeled 43.3% of patients as high-risk and obtained a PPV of 15.1% and an NPV of 98.5%.

**Figure 4:**
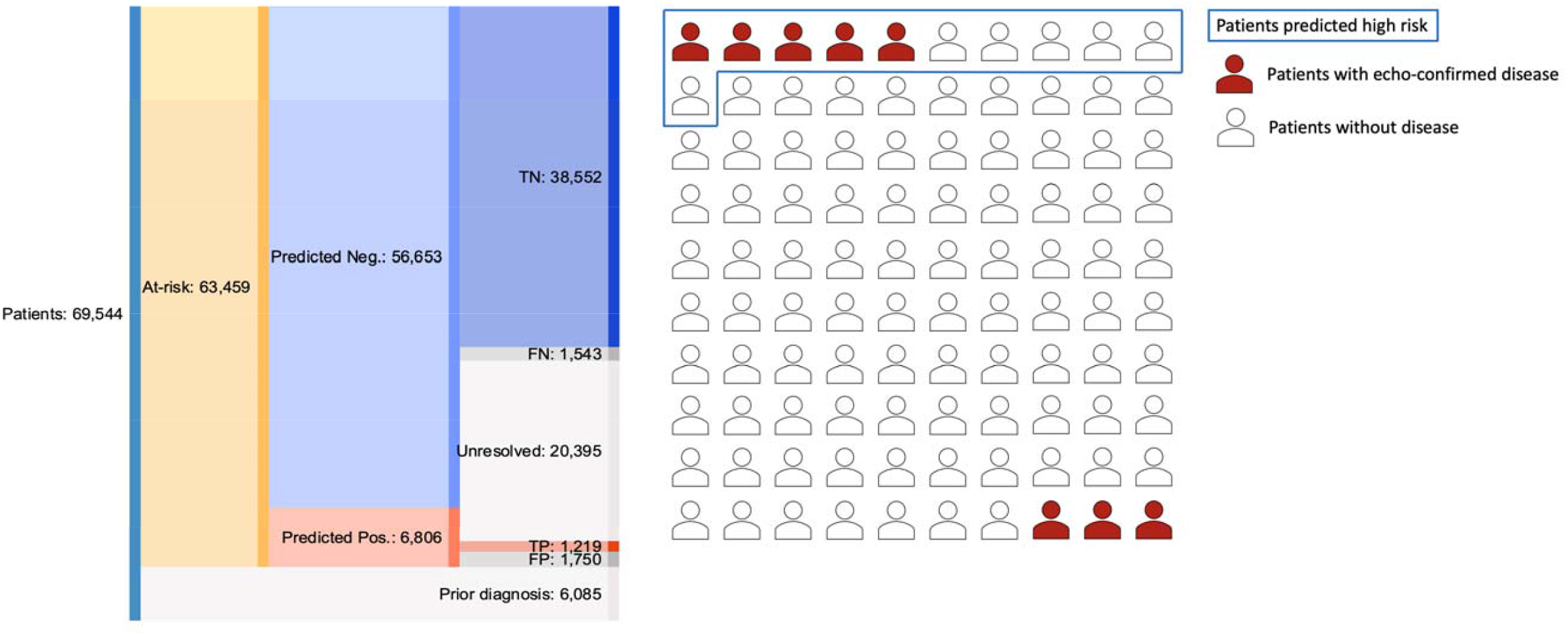
Results of retrospective deployment scenario from 2010 for (A) all patients and (B) relative results per 100 at-risk patients. These results are based on a threshold yielding 50% sensitivity from the pre-2010 cross-validation experiment, resulting in 41.1% PPV, 96.2% NPV, 95.7% specificity, 44.1% sensitivity, and 6.4% prevalence in 2010. For 100 patients without known history of disease obtaining an ECG, the rECHOmmend model will identify 11 patients at high-risk of disease, of which 5 are expected to have true disease within 1 year. The model will identify 89 patients not at high-risk of disease, of which 86 are not expected to have true disease within 1 year.

For a more practical comparison, using a threshold estimated to yield 50% sensitivity, the deployment model labeled 10.7% of patients as high-risk for any of the seven disease outcomes. Among 2969 predicted high-risk patients with adequate follow-up who met our definition for the composite label, 1219 patients were diagnosed with at least one of the disease outcomes within a year, a PPV of 41.1%. Of these 1219 patients, 137 (11%) received a diagnosis of AS, 86 (7%) of AR, 387 (32%) of MR, 17 (1%) of MS, 375 (31%) of TR, 785 (64%) of reduced EF, and 280 (23%) of IVS thickening. Among 40,095 predicted low-risk patients with adequate follow-up and defined labels, 38,552 patients did not develop any of the outcomes within a year, a NPV of 96.2%.

Overall, at this model threshold, for every 100 at-risk patients who obtained an ECG, our model would identify 11 as high-risk, of which 5 would truly have echocardiography-confirmed disease, and 89 as low-risk, of which 86 would truly not have disease within 1 year (Figure 4B).

### Sensitivity Analyses

When using severe-only disease labels, AUROCs across input feature combinations for the composite endpoint were similar to the primary results (Table 2), ranging from 0.79 for age and sex only to 0.94 for all inputs (Supplemental Table 11). AUPRC and PPV at 90% sensitivity were lower given the lower prevalence of severe-only disease. Across the individual diseases, AUROC of the age, sex, and ECG traces model was again similar, ranging from 0.84-0.96, and again with lower AUPRC and PPV due to the lower prevalence (Supplemental Table 12). The overall rECHOmmend model using severe-only disease labels attained an AUROC of 0.92 with a PPV of 31.2% at 90% sensitivity with 10.6% disease prevalence.

When using echocardiography-confirmed labels only, AUROC was slightly lower than our primary results, while AUPRC and PPV at 90% sensitivity was higher (Supplemental Tables 13-14). This was likely due to the artificially higher prevalence, as the number of negative patients decreased with this requirement for echocardiography-confirmed absence of disease. The overall rECHOmmend model obtained an AUROC of 0.88 with a 74% PPV at 90% sensitivity with 53% disease prevalence.

## Discussion

We developed a machine-learning platform called “rECHOmmend,” which can predict clinically significant valvular disease, reduced left ventricular EF, or pathologically increased septal thickness with excellent performance (AUROC 0.91) by using only ECG traces, age, and sex. Furthermore, we demonstrated that the combination of these distinct endpoints into a single platform tied to a recommendation for a singular, practical clinical response—follow-up echocardiography—resulted in an overall PPV of 42% for clinically meaningful disease while maintaining high sensitivity (90%) and specificity (73%). This suggests that for the millions of patients who receive an ECG each year without pre-existing structural heart disease, nearly half of patients deemed high-risk by this model would be found to have true disease within a year. We confirmed the validity of this approach through a multi-site validation on non-overlapping data sets from multiple clinical sites across the Geisinger system. Moreover, we confirmed the clinical utility of this approach in our retrospective deployment, as our model trained on pre-2010 data and deployed on all patients without prior disease who obtained an ECG in 2010 maintained similarly high performance as compared to the main cross-validation results based only on passive observation and standard clinical care. With an active deployment of the rECHOmmend platform, even higher yields / PPV are anticipated once clinicians can pursue active intervention in the form of follow-up echocardiogram or more detailed history-taking and physical examination.

Clinically, this model enables targeted echocardiographic screening to help detect unrecognized and underdiagnosed diseases. Currently echocardiography is not used for population screening given the low prevalence of disease in the general population, as prior attempts were shown to be ineffective.^22,23^ Therefore, indicated use of echocardiography is typically triggered by a symptom, adverse event, physical exam, or incidental finding leading to suspicion of heart disease, raising the pretest probability and likelihood of a clinically impactful or actionable finding.^6,7,24^ However, a significant gap remains in that a large number of patients, in meeting that triggered indication for suspected disease, will have already suffered an adverse event, a symptom affecting their quality of life, or an irreversible pathophysiologic change from their undiagnosed disease. For example, in severe AS, the initial presenting symptom is reduced EF for 8% of patients, angina for 35-41%, and syncope for 10-11% of patients, which may lead to falls, hip fractures, or reduced functional status.^25–27^ Prior studies have also shown that up to half of elderly patients have undiagnosed valvular disease, including 11.3% with moderate or severe disease, while the majority of patients with hypertrophic cardiomyopathy may be undiagnosed, and nearly 50% of patients with EF <40% are asymptomatic.^11,12,28^ This rECHOmmend model, with both high sensitivity and precision, can help guide the decision to obtain an echocardiogram even for asymptomatic patients, shifting the balance to a scenario where echocardiography can be an effective screening tool to help clinicians diagnose patients at the right time to prevent downstream adverse events, optimize the timing of interventions, and better implement evidence-based monitoring or management.

Our findings also suggest a path toward overcoming some of the existing challenges with clinical implementation of ECG prediction models. This novel approach of combining multiple endpoints which align under the same recommended clinical action enables the model to leverage the increased prevalence and probability of any one disease state occurring to improve predictive performance for potential clinical implementation. Previous studies have shown that CNN-based ECG prediction models can predict a variety of cardiovascular outcomes including atrial fibrillation, aortic stenosis, and LV dysfunction with good performance, with AUROCs from 0.80-0.93.^13–16,29^ However, concerns often exist around real-world implementation of such models due to limitations in precision and recall, concerns regarding the negative impact of false positives, and limited actionability or portability.^30^ Our models compare favorably to those in the literature, with similar or higher AUROCs and higher precision or PPV, but also result in a clearly actionable recommendation while remaining highly portable. Our featured model results of 0.91 AUROC, 42% PPV and 90% sensitivity on cross-validation is based on age, sex, and ECG traces alone as inputs, which we believe represents the optimal balance between performance and portability, While the addition of EHR data did slightly improve performance, there would be a major tradeoff in decreased portability with the need for EHR or clinical data warehouse integration. This model uses data readily available from any ECG system, such as MUSE, and could be easily deployed across most healthcare systems.

We also find that simulated deployment on large retrospective datasets can shed light on important questions and estimate true clinical impact prior to the costly implementation of prediction models in practice or clinical trials, where performance may differ from strictly cross-validation performance of the same models.^13,31^ In our simulated deployment on ECGs from 2010, 11% of at-risk patients without history of disease were predicted to be high-risk, of which 41% with adequate follow-up were truly diagnosed with disease in the following year, through only standard clinical care and without any clinician behavior change or active intervention that true deployment may elicit. This suggests that this 41% PPV is likely a lower bound for the expected real-world performance of the rECHOmmend model. Our simulated real-world deployment scenario compares favorably with a recent pragmatic trial for predicting reduced EF which identified a real-world PPV of 39% using an EF cutoff of <=50%, of which 24% of patients meeting this definition qualified with an EF of exactly 50%.^31^ Deployment scenarios also demonstrate that cross-validation metrics which depend on prevalence likely overestimate real-world performance as seen in recent studies, including for the above reduced EF trial which lagged behind the original cross-validation results (reported PPV of 63%).^15,31^ We propose that simulated retrospective deployment be carried out for future prediction models to better gauge feasibility and real-world performance prior to clinical implementation.

Our study has several limitations. Training and evaluation were limited to a regional health system where most patients are white, so results may not be generalizable to hospitals or regions with more diversity. We are not aware of any physiologic differences across race/ethnicity that would lead these ECG-based models to perform differently across groups, corroborated by prior studies,^32^ but results should be confirmed in further research. In addition, we used echocardiography-confirmed diagnoses to generate our positive labels, which were confirmed on chart review to have a high PPV but there may be additional patients with disease—false negatives—who were not captured using this method. However, given the low prevalence of each disease in the general population and echocardiography being the diagnostic standard, the negatives are likely true negatives, as seen in the retrospective deployment where we leveraged up to a decade of follow-up to determine negative outcomes. In addition, this machine-learning approach has limited interpretability in identifying feature importance. Finally, increased IVS thickness may represent infiltrative diseases, hypertrophic cardiomyopathy, or may largely represent concentric remodeling related to longstanding, poorly controlled hypertension; however, these conditions are all clinically actionable.

This study demonstrates that a machine-learning model using only ECG-based inputs can predict multiple important cardiac endpoints within a single platform with both good performance and high PPV, thereby representing a practical tool with which to better target echocardiography to detect undiagnosed disease. We confirmed these results through retrospective real-world deployment scenarios to show the large impact that such a model can have on patients when deployed across a health system. These approaches to both clinical predictions and simulated deployment represent practical solutions for existing limitations in the implementation of machine learning in healthcare, hopefully bringing this technology one step closer to standard clinical practice.

## Data Availability

All intermediate, subgroup, and aggregate results are publicly available online as a searchable dashboard. Patient-level data are not available for the Geisinger data set. Requests for code or data can be made to the corresponding author.

http://rechommend.herokuapp.com/

## Declaration of interests

Geisinger investigators (AUC, LJ, SR, JAR, DBR, JBL, CMH, RC) receive funding from Tempus for ongoing development of predictive modeling technology. Tempus and Geisinger have jointly applied for predictive modeling patents. None of the Geisinger investigators have ownership interest in any of the intellectual property resulting from the partnership. Tempus did not have any input in the design, execution, interpretation of results or decision to publish. JMP, NZ, GL, and BFK are Tempus employees. SRS is a consultant for Tempus. SRS is also an employee of physIQ and reports personal fees from Otsuka and Janssen, outside the submitted work. BKF reports personal fees from Novartis, outside the submitted work.

## Acknowledgements

This work is supported by a grant from Tempus.

## Author Contributions

AUC, LJ, JMP, SR, DBR, CMH, BKF, and RC contributed to study design, implementation, execution, data analysis, interpretation, and paper writing. JAR, JBL, GL, NZ, SRS, and CWG contributed to study design, data interpretation, and paper writing.

## Notes

### Author Declarations

The Institutional Review Board of Geisinger approved this study with a waiver of consent

