## Supplement for "rECHOmmend: an ECG-based machine-learning approach for identifying patients at high-risk of undiagnosed structural heart disease detectable by echocardiography"

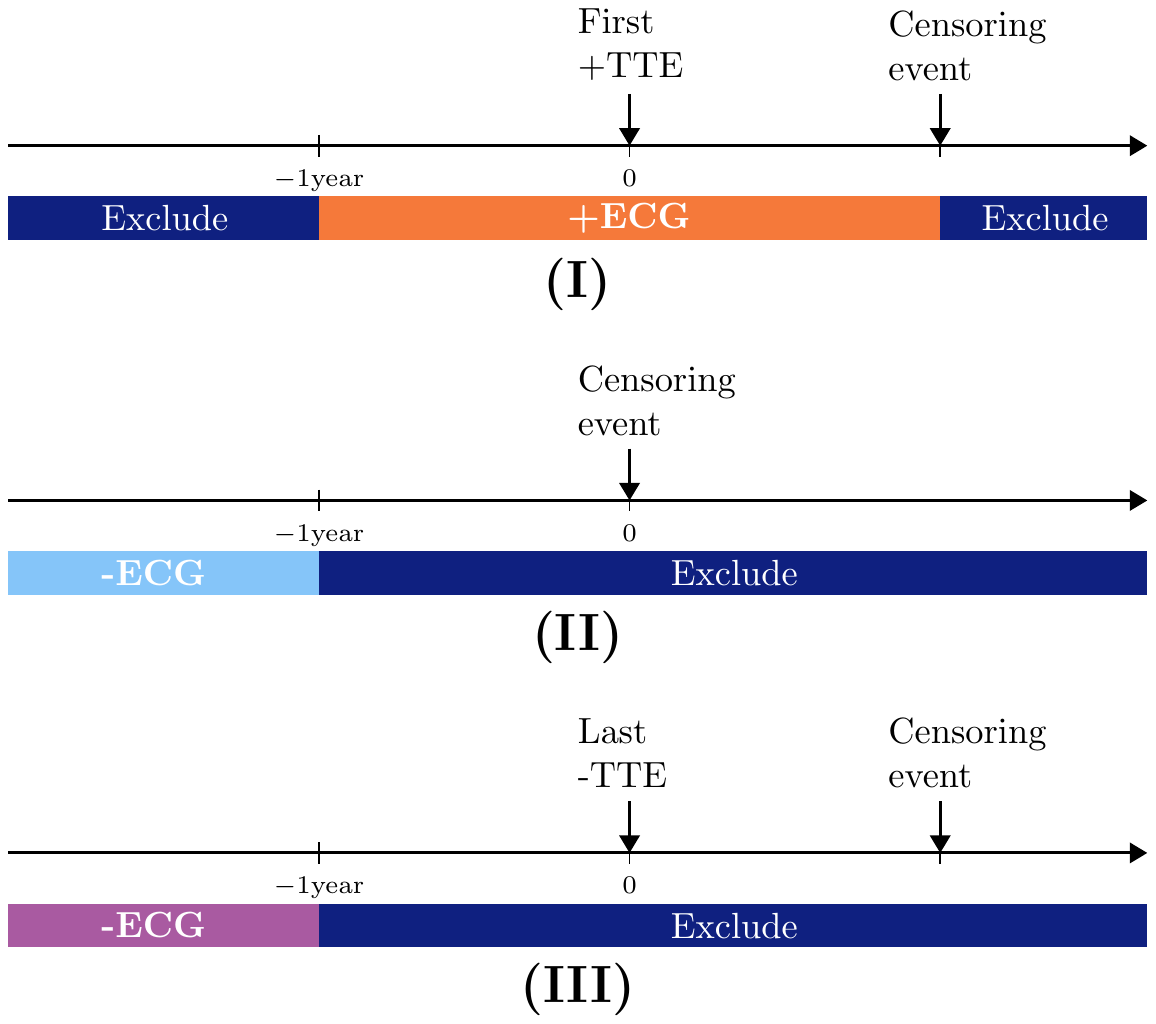

Supplemental Figure 1: (A) Patient timeline used to label (I) positive ECGs (+ECG in orange background) and negative ECGs as either (II) or (III). For our sensitivity analysis, we also defined (III) echo-confirmed negative ECGs. Censoring events are any intervention that could modify the underlying physiology of the disease of interest such as a valve replacement or the end of observation/follow-up in the EHR. ‘Last negative TTE’ requires that no record of a prior positive TTE exists.

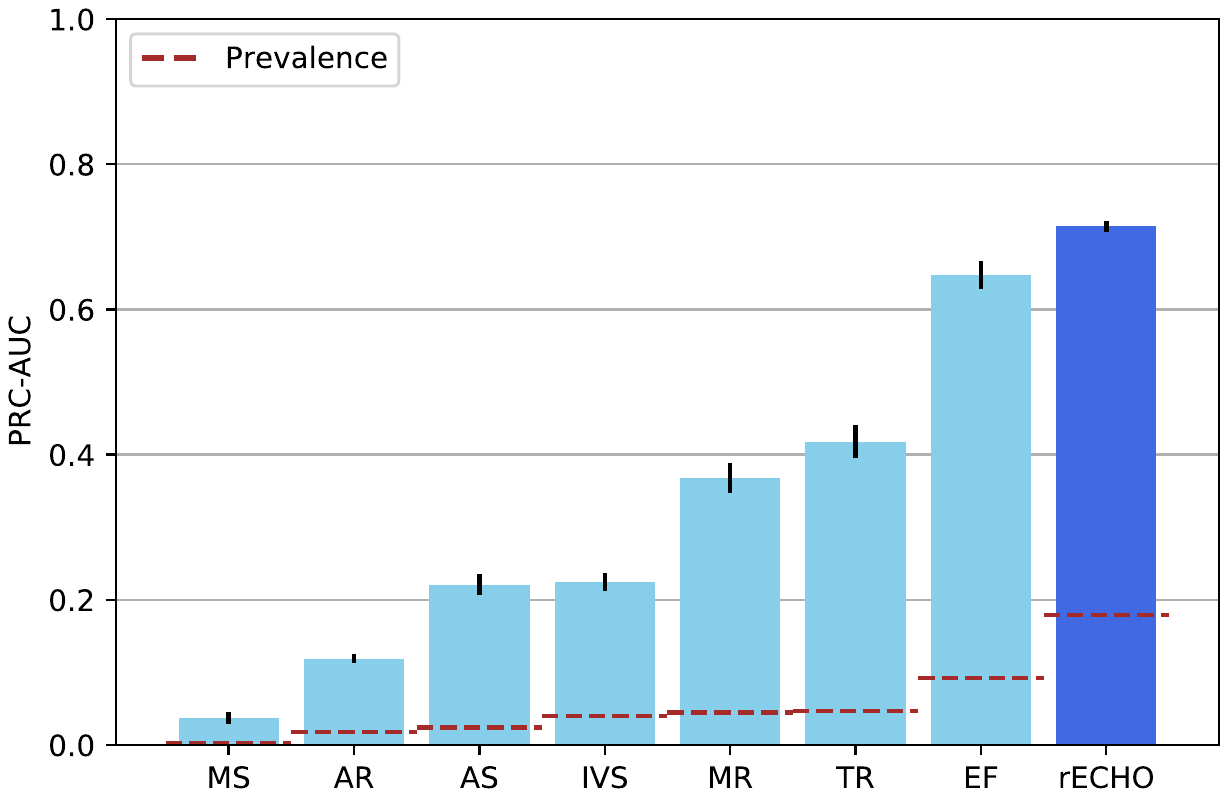

Supplemental Figure 2: Area under the Precision-Recall curve (AUPRC) for each of the individual diseases and the rECHOmmend model. The red dashed line shows the prevalence for each of the labels

**Supplemental Table 1:** List of inputs grouped by category. Each input is shown with its units in parenthesis. The structured ECG findings were all binary true/false variables.

|  | List of inputs |
| --- | --- |
| Demographics and Vitals | Age (years), race (white/other), sex, smoke (ever), BMI (kg/m2), diastolic and systolic blood pressure (mmHg), heart rate (bpm), height (cm), weight (kg). |
| Labs | A1C (%), Bilirubin (mg/dl), BUN (mg/dl), Cholesterol (mg/dl), CKMB (ng/ml), Creatinine (mg/dl), CRP (mg/l), D dimer (mcg/ml FEU), Glucose (mg/dl), HDL (mg/dl), Hemoglobin (g/dl), LDH (u/l), LDL (mg/dl), Lymphocytes (%), Potassium (mmol/l), PRO BNP (pg/ml), Sodium (mmol/l), Troponin I and T (ng/ml), Triglyceride (mg/dl), Uric acid (mg/dl), VLDL (mg/dl), eGFR (ml/min/1.73m^2^) |
| ECG findings | acute myocardial infarction, atrial fibrillation, atrial flutter, complete block, early repolarization, fascicular block, first-degree atrioventricular block, intraventricular block, incomplete left BBB, incomplete right BBB, ischemia, left axis deviation, left BBB, low QRS voltage, left ventricular hypertrophy, nonspecific ST abnormality, nonspecific T-wave abnormality, normal, other bradycardia, premature atrial contractions, pacemaker, poor tracing, prior infarct, prior MI anterior, prolonged QT, premature ventricular contractions, right axis deviation, right BBB, second-degree atrioventricular block, sinus bradycardia, supraventricular tachycardia, tachycardia, T inversion, ventricular tachycardia |
| ECG measurements | Average RR interval (ms), PR interval (ms), P axis (degrees), QRS duration (ms), QT (ms), QTC (ms), R axis (degrees), T axis (degrees), Ventricular rate (bpm) |

**Supplemental Table 2**: Key words and phrases for assigning an abnormality and severity to each valve in an echocardiography report. The search is not case sensitive.

| **Abnormality** | |
| --- | --- |
| Stenosis | stenosis, stenotic |
| Regurgitation | regurgitation, regurgitant, insufficiency |
| **Severity** | |
| Normal or Mild (negative TTE) | absent, no stenosis, no AS, no MS, not stenotic, no PS, no tricuspid stenosis, no significant, no regurgitation, No TR, No MR, TS excluded, MS excluded, AS excluded, w/o stenosis, no mitral, no AR, trace, no evidence of, no pulmonic, no mitral, without aortic stenosis, stenosis is absent, no mitral regurgitation, physiologic, no hemodynamically, Normal 2-D, Normal MV, not sign, Normal structure and function, normal prosthetic, normal function, function normal, There is a normal amount of, is probably normal, is normal without, mild, valvular, aortic stenosis is present, valve stenosis is present, stenosis is possible, stenosis is possibly present, borderline |
| Moderate | moderate, mod |
| Severe | severe, possibly, severe, moderate-severe, mod-severe, moderate - severe, moderately severe, moderate to severe, critical, consistent with significant |
| **Valve** | |
| Aortic | aortic, AS, AR, AV |
| Tricuspid | tricuspid, TR, TS, TV |
| Mitral | mitral, MR, MS, MV |

**Supplemental Table 3**: List of ICD 10 and ICD 9 codes used to define relevant disease within the EHR. Used to define negative disease labels for ECGs of patients without history of echocardiography and adequate follow-up, if the patient also did not have any of these codes for each given disease label.

| Diagnosis | ICD 10 Codes | ICD 9 Codes |
| --- | --- | --- |
| AR | I06.1, I06.2, I06.8, I06.9, I08.0, I08.2, I08.3, I08.8, I08.9, I33.0, I33.9, I35.1, I35.2, I35.8, I35.9, Q20.0, Q20.1, Q20.2, Q20.3, Q20.4, Q20.5, Q20.6, Q20.8, Q20.9, Q21.0, Q21.1, Q21.2, Q21.3, Q21.4, Q21.8, Q21.9, Q22.0, Q22.1, Q22.2, Q22.3, Q22.4, Q22.5, Q22.6, Q22.8, Q22.9, Q23.0, Q23.1, Q23.2, Q23.3, Q23.4, Q23.8, Q23.9, Q24.0, Q24.1, Q24.2, Q24.3, Q24.4, Q24.5, Q24.6, Q24.8, Q24.9, Z95.4 | 395.1, 395.2, 395.9, 396.0, 396.1, 396.2, 396.3, 396.8, 396.9, 397.9, 421.0, 421.9, 424.1, 745.0, 745.10, 745.11, 745.12, 745.19, 745.2, 745.3, 745.4, 745.5, 745.60, 745.61, 745.69, 745.7, 745.8, 745.9, 746.00, 746.01, 746.02, 746.09, 746.1, 746.2, 746.3, 746.4, 746.5, 746.6, 746.7, 746.81, 746.82, 746.83, 746.84, 746.85, 746.86, 746.87, 746.89, 746.9, V42.2 |
| AS | I06.0, I06.2, I06.8, I06.9, I08.0, I08.2, I08.3, I08.8, I08.9, I33.0, I33.9, I35.0, I35.2, I35.8, I35.9, Q20.0, Q20.1, Q20.2, Q20.3, Q20.4, Q20.5, Q20.6, Q20.8, Q20.9, Q21.0, Q21.1, Q21.2, Q21.3, Q21.4, Q21.8, Q21.9, Q22.0, Q22.1, Q22.2, Q22.3, Q22.4, Q22.5, Q22.6, Q22.8, Q22.9, Q23.0, Q23.1, Q23.2, Q23.3, Q23.4, Q23.8, Q23.9, Q24.0, Q24.1, Q24.2, Q24.3, Q24.4, Q24.5, Q24.6, Q24.8, Q24.9, Q25.3, Z95.4 | 395.0, 395.2, 395.9, 396.0, 396.1, 396.2, 396.3, 396.8, 396.9, 397.9, 421.0, 421.9, 424.1, 745.0, 745.10, 745.11, 745.12, 745.19, 745.2, 745.3, 745.4, 745.5, 745.60, 745.61, 745.69, 745.7, 745.8, 745.9, 746.00, 746.01, 746.02, 746.09, 746.1, 746.2, 746.3, 746.4, 746.5, 746.6, 746.7, 746.81, 746.82, 746.83, 746.84, 746.85, 746.86, 746.87, 746.89, 746.9, 747.22, V42.2 |
| MR | I05.1, I05.2, I05.8, I05.9, I08.0, I08.1, I08.3, I08.8, I08.9, I33.0, I33.9, I34.0, I34.1, I34.8, I34.9, Q20.0, Q20.1, Q20.2, Q20.3, Q20.4, Q20.5, Q20.6, Q20.8, Q20.9, Q21.0, Q21.1, Q21.2, Q21.3, Q21.4, Q21.8, Q21.9, Q22.0, Q22.1, Q22.2, Q22.3, Q22.4, Q22.5, Q22.6, Q22.8, Q22.9, Q23.0, Q23.1, Q23.2, Q23.3, Q23.4, Q23.8, Q23.9, Q24.0, Q24.1, Q24.2, Q24.3, Q24.4, Q24.5, Q24.6, Q24.8, Q24.9, Z95.4 | 394.1, 394.2, 394.9, 396.0, 396.1, 396.2, 396.3, 396.8, 396.9, 397.9, 421.0, 421.9, 424.0, 745.0, 745.10, 745.11, 745.12, 745.19, 745.2, 745.3, 745.4, 745.5, 745.60, 745.61, 745.69, 745.7, 745.8, 745.9, 746.00, 746.01, 746.02, 746.09, 746.1, 746.2, 746.3, 746.4, 746.5, 746.6, 746.7, 746.81, 746.82, 746.83, 746.84, 746.85, 746.86, 746.87, 746.89, 746.9, V42.2 |
| MS | I05.0, I05.2, I05.8, I05.9, I08.0, I08.1, I08.3, I08.8, I08.9, I33.0, I33.9, I34.2, I34.8, I34.9, Q20.0, Q20.1, Q20.2, Q20.3, Q20.4, Q20.5, Q20.6, Q20.8, Q20.9, Q21.0, Q21.1, Q21.2, Q21.3, Q21.4, Q21.8, Q21.9, Q22.0, Q22.1, Q22.2, Q22.3, Q22.4, Q22.5, Q22.6, Q22.8, Q22.9, Q23.0, Q23.1, Q23.2, Q23.3, Q23.4, Q23.8, Q23.9, Q24.0, Q24.1, Q24.2, Q24.3, Q24.4, Q24.5, Q24.6, Q24.8, Q24.9, Z95.4 | 394.0, 394.2, 394.9, 396.0, 396.1, 396.2, 396.3, 396.8, 396.9, 397.9, 421.0, 421.9, 424.0, 745.0, 745.10, 745.11, 745.12, 745.19, 745.2, 745.3, 745.4, 745.5, 745.60, 745.61, 745.69, 745.7, 745.8, 745.9, 746.00, 746.01, 746.02, 746.09, 746.1, 746.2, 746.3, 746.4, 746.5, 746.6, 746.7, 746.81, 746.82, 746.83, 746.84, 746.85, 746.86, 746.87, 746.89, 746.9, V42.2 |
| TR | I07.1, I07.2, I07.8, I07.9, I08.1, I08.2, I08.3, I08.8, I08.9, I33.0, I33.9, I36.1, I36.2, I36.8, I36.9, Q20.0, Q20.1, Q20.2, Q20.3, Q20.4, Q20.5, Q20.6, Q20.8, Q20.9, Q21.0, Q21.1, Q21.2, Q21.3, Q21.4, Q21.8, Q21.9, Q22.0, Q22.1, Q22.2, Q22.3, Q22.4, Q22.5, Q22.6, Q22.8, Q22.9, Q23.0, Q23.1, Q23.2, Q23.3, Q23.4, Q23.8, Q23.9, Q24.0, Q24.1, Q24.2, Q24.3, Q24.4, Q24.5, Q24.6, Q24.8, Q24.9, Z95.4 | 396.8, 396.9, 397.0, 397.9, 421.0, 421.9, 424.2, 745.0, 745.10, 745.11, 745.12, 745.19, 745.2, 745.3, 745.4, 745.5, 745.60, 745.61, 745.69, 745.7, 745.8, 745.9, 746.00, 746.01, 746.02, 746.09, 746.1, 746.2, 746.3, 746.4, 746.5, 746.6, 746.7, 746.81, 746.82, 746.83, 746.84, 746.85, 746.86, 746.87, 746.89, 746.9, V42.2 |
| EF < 50% | B33.2, I09.81, I25.5, I42.0, I42.6, I42.7, I42.8, I42.9, I50.1, I50.2, I50.20, I50.21, I50.22, I50.23, I50.3, I50.30, I50.31, I50.32, I50.33, I50.4, I50.40, I50.41, I50.42, I50.43, I50.8, I50.81, I50.810, I50.811, I50.812, I50.813, I50.814, I50.82, I50.83, I50.84, I50.89, I50.9, I51.8, I51.81, I51.89, I97.13, O90.3, Q20.0, Q20.1, Q20.2, Q20.3, Q20.4, Q20.5, Q20.6, Q20.8, Q20.9, Q21.0, Q21.1, Q21.2, Q21.3, Q21.4, Q21.8, Q21.9, Q22.0, Q22.1, Q22.2, Q22.3, Q22.4, Q22.5, Q22.6, Q22.8, Q22.9, Q23.0, Q23.1, Q23.2, Q23.3, Q23.4, Q23.8, Q23.9, Q24.0, Q24.1, Q24.2, Q24.3, Q24.4, Q24.5, Q24.6, Q24.8, Q24.9, T86.2, T86.3, Z94.1, Z94.3 | 398.91, 414.8, 425.2, 425.4, 425.5, 425.9, 428.0, 428.1, 428.20, 428.21, 428.22, 428.23, 428.30, 428.31, 428.32, 428.33, 428.40, 428.41, 428.42, 428.43, 428.9, 429.82, 429.83, 429.89, 674.52, 674.54, 745.0, 745.10, 745.11, 745.12, 745.19, 745.2, 745.3, 745.4, 745.5, 745.60, 745.61, 745.69, 745.7, 745.8, 745.9, 746.00, 746.01, 746.02, 746.09, 746.1, 746.2, 746.3, 746.4, 746.5, 746.6, 746.7, 746.81, 746.82, 746.83, 746.84, 746.85, 746.86, 746.87, 746.89, 746.9, V42.1 |
| IVS>15mm | D86.0, D86.1, D86.2, D86.3, D86.8, D86.81, D86.82, D86.83, D86.84, D86.85, D86.86, D86.87, D86.89, D86.9, E74.0, E74.00, E74.01, E74.02, E74.03, E74.04, E74.09, E74.1, E74.10, E74.11, E74.12, E74.19, E74.2, E74.20, E74.21, E74.29, E74.3, E74.31, E74.39, E74.4, E74.8, E74.810, E74.818, E74.819, E74.89, E74.9, E75.0, E75.00, E75.01, E75.02, E75.09, E75.1, E75.10, E75.11, E75.19, E75.2, E75.21, E75.22, E75.23, E75.24, E75.240, E75.241, E75.242, E75.243, E75.248, E75.249, E75.25, E75.26, E75.29, E75.3, E75.4, E75.5, E75.6, E83.11, E85.0, E85.1, E85.2, E85.3, E85.4, E85.8, E85.81, E85.82, E85.89, E85.9, I11.0, I11.9, I12.0, I12.9, I13.0, I13.1, I13.10, I13.11, I13.2, I15.0, I15.1, I15.2, I15.8, I15.9, I16.0, I16.1, I16.9, I37.1, I37.2, I42.1, I42.2, Q20.0, Q20.1, Q20.2, Q20.3, Q20.4, Q20.5, Q20.6, Q20.8, Q20.9, Q21.0, Q21.1, Q21.2, Q21.3, Q21.4, Q21.8, Q21.9, Q22.0, Q22.1, Q22.2, Q22.3, Q22.4, Q22.5, Q22.6, Q22.8, Q22.9, Q23.0, Q23.1, Q23.2, Q23.3, Q23.4, Q23.8, Q23.9, Q24.0, Q24.1, Q24.2, Q24.3, Q24.4, Q24.5, Q24.6, Q24.8, Q24.9, Q25.0, Q25.1, Q25.2, Q25.21, Q25.29, Q25.3, Q25.4, Q25.40, Q25.41, Q25.42, Q25.43, Q25.44, Q25.45, Q25.46, Q25.47, Q25.48, Q25.49, Q25.5, Q25.6, Q25.7, Q25.71, Q25.72, Q25.79, Q25.8, Q25.9, T86.2, T86.3, Z94.1, Z94.3 | 271.0, 271.1, 271.2, 271.3, 271.4, 271.8, 271.9, 272.7, 272.8, 272.9, 277.30, 277.31, 277.39, 330.0, 330.1, 401.0, 401.1, 401.9, 401.9, 402.0, 402.00, 402.00, 402.01, 402.01, 402.1, 402.10, 402.10, 402.11, 402.11, 402.9, 402.90, 402.90, 402.91, 402.91, 403.0, 403.00, 403.00, 403.01, 403.01, 403.1, 403.10, 403.10, 403.11, 403.11, 403.9, 403.90, 403.90, 403.91, 403.91, 404.0, 404.00, 404.00, 404.01, 404.01, 404.02, 404.02, 404.03, 404.03, 404.1, 404.10, 404.10, 404.11, 404.11, 404.12, 404.12, 404.13, 404.13, 404.9, 404.90, 404.90, 404.91, 404.91, 404.92, 404.92, 404.93, 404.93, 405.0, 405.01, 405.01, 405.09, 405.09, 405.1, 405.11, 405.11, 405.19, 405.19, 405.9, 405.91, 405.91, 405.99, 405.99, 425.11, 425.18, 745.0, 745.10, 745.11, 745.12, 745.19, 745.2, 745.3, 745.4, 745.5, 745.60, 745.61, 745.69, 745.7, 745.8, 745.9, 746.00, 746.01, 746.02, 746.09, 746.1, 746.2, 746.3, 746.4, 746.5, 746.6, 746.7, 746.81, 746.82, 746.83, 746.84, 746.85, 746.86, 746.87, 746.89, 746.9, 747.0, 747.10, 747.11, 747.20, 747.21, 747.22, 747.29, 747.31, 747.32, 747.39, V42.1 |

**Supplemental Table 4**: Single output low-parameter convolutional neural network (CNN) design for training on 8 non-derived ECG leads. The network contains a total of 18,945 trainable and 384 non-trainable parameters. Both Dropout layers were set at 25% drop rate. CBR is a brief notation for a sequence of 1D CNN, batch normalization, and ReLU layers.

| **Layer** | **Output Shape** | **#Parameters** |
| --- | --- | --- |
| Input | (5000,8) | 0 |
| Rescaling | (5000,8) | 0 |
| CBR-1 | (5000,16) | 656+64 |
| CBR-2 | (5000,16) | 1,296+64 |
| MaxPool1D | (1666,16) | 0 |
| CBR-3 | (1666,16) | 1,296+64 |
| CBR-4 | (1666,16) | 1,296+64 |
| MaxPool1D | (555,16) | 0 |
| CBR-5 | (555,16) | 1,296+64 |
| CBR-6 | (555,16) | 1,296+64 |
| MaxPool1D | (185,16) | 0 |
| CBR-7 | (185,16) | 1,296+64 |
| CBR-8 | (185,16) | 1,296+64 |
| MaxPool1D | (61,16) | 0 |
| CBR-9 | (61,16) | 1,296+64 |
| CBR-10 | (61,16) | 1,296+64 |
| MaxPool1D | (20,16) | 0 |
| CBR-11 | (20,16) | 1,296+64 |
| CBR-12 | (20,16) | 1,296+64 |
| MaxPool1D | (6,16) | 0 |
| Flatten | (96,) | 0 |
| Dense + Dropout | (32,) | 3104 |
| Dense + Dropout | (16,) | 528 |
| Dense | (1,) | 17 |

**Supplemental Table 5**: Echo label count and relative prevalence for each diagnosis among total 758,269 echocardiograms.

|  | **Normal-Mild  (Negative)** | **Moderate-Severe  (Positive)** | **Prevalence** |
| --- | --- | --- | --- |
| Aortic Stenosis | 436,886 | 27,857 | 6.0% |
| Aortic Regurgitation | 443,060 | 19,808 | 4.3% |
| Mitral regurgitation | 453,554 | 43,280 | 8.7% |
| Mitral Stenosis | 493,465 | 2,908 | 0.6% |
| Tricuspid Regurgitation | 437,418 | 44,533 | 9.2% |
|  | **False  (Negative)** | **True  (Positive)** |  |
| Ejection Fraction<50% | 459,281 | 95,331 | 17.2% |
| IVS thickness>15mm | 490,830 | 32,593 | 6.2% |

**Supplemental Table 6**: Count of ECGs and total prevalence for each diagnosis among 2,232,130 ECGs with at least one disease label, based on the positive and negative label definitions outlined in the Methods.

|  | **Negative** | **Positive** | **Prevalence** | **Undefined** |
| --- | --- | --- | --- | --- |
| Aortic Stenosis | 1,695,186 | 64,625 | 2.9% | 472,319 |
| Aortic Regurgitation | 1,706,295 | 45,333 | 2.0% | 480,502 |
| Mitral Regurgitation | 1,606,731 | 142,209 | 6.4% | 483,190 |
| Mitral Stenosis | 1,743,273 | 8,034 | 0.4% | 480,823 |
| Tricuspid Regurgitation | 1,647,485 | 151,971 | 6.8% | 432,674 |
| Ejection Fraction<50% | 1,416,373 | 322,482 | 14.4% | 493,275 |
| IVS thickness>15mm | 1,643,697 | 123,676 | 5.5% | 464,757 |
| rECHOmmend | 1,075,298 | 576,654 | 25.8% | 580,178 |

**Supplemental Table 7**: Average value for each predictor grouped by rECHOmmend labeled ECGs. Negative refers to ECGs from patients that were not diagnosed with any of the 7 diseases within a year. Positive refers to ECGs from patients that were diagnosed with at least one of the 7 diseases within a year or before the ECG acquisition time. Unresolved refers to ECGs that did not meet the composite criteria. Baseline characteristics of patients with defined vs undefined labels in a random ECG per patient for ECGs with at least a label. We show mean (SD) for continuous and percentage for binary features.

|  | **Negative**  **N=1,075,298** | **Positive**  **N=576,654** | **Defined**  **N=1,651,952** | **Undefined**  **N=1,273,973** |
| --- | --- | --- | --- | --- |
| Race (%White) | 96.6% | 97.4% | 97.10% | 96.4% |
| Sex (%Male) | 45.9% | 58.4% | 50.10% | 50.5% |
| Smoker (%Ever) | 57.6% | 63.0% | 59.70% | 58.7% |
| Age (years) | 57 (17) | 71 (14) | 63 (17) | 64 (17) |
| BMI (kg/m2) | 31 (8) | 30 (9) | 31 (9) | 31 (9) |
| Diastolic BP (mmHg) | 75 (11) | 70 (13) | 73 (12) | 73 (12) |
| Systolic BP (mmHg) | 129 (19) | 128 (21) | 129 (20) | 129 (20) |
| Heart Rate (bpm) | 77 (15) | 76 (17) | 76 (15) | 77 (16) |
| Height (cm) | 168 (11) | 169 (11) | 168 (11) | 169 (11) |
| Weight (kg) | 88 (24) | 86 (25) | 88 (24) | 87 (25) |
| A1C | 6.8 (4.8) | 6.9 (1.6) | 6.9 (3) | 6.8 (1.6) |
| Bilirubin | 0.5 (0.6) | 0.6 (0.7) | 0.57 (0.60) | 0.6 (0.9) |
| BUN | 17 (9) | 26 (16) | 20.5 (12.8) | 20 (12) |
| Cholesterol | 184 (45) | 159 (47) | 172 (47) | 171 (48) |
| CKMB | 7 (24) | 10 (37) | 8.9 (32.2) | 7 (25) |
| Creatinine | 1.0 (1.7) | 1.4 (1.3) | 1.2 (1.4) | 1.1 (1.0) |
| CRP | 24 (52) | 49 (71) | 36.2 (63.9) | 36 (63) |
| D dimer | 1.0 (2.0) | 2.0 (3.1) | 1.5 (2.6) | 1.5 (2.8) |
| Glucose | 115 (44) | 124 (53) | 119 (48) | 120 (49) |
| HDL | 50 (16) | 46 (16) | 48 (16) | 48 (16) |
| Hemoglobin | 14 (20) | 14 (44) | 14 (34) | 13 (27) |
| LDH | 219 (134) | 273 (288) | 249 (237) | 264 (256) |
| LDL | 104 (37) | 86 (37) | 95 (38) | 94 (38) |
| Lymphocytes | 25 (10) | 20 (11) | 23 (11) | 22 (11) |
| Potassium | 4.2 (0.8) | 4.3 (0.7) | 4.2 (0.7) | 4.2 (0.7) |
| Pro-BNP | 893 (3399) | 6881 (12339) | 5002 (10668) | 2655 (7653) |
| Sodium | 139 (3) | 139 (4) | 139 (3) | 139 (4) |
| Troponin I | 0.8 (11.9) | 1.1 (10.2) | 1 (13) | 0.8 (11.7) |
| Troponin T | 0.1 (0.4) | 0.2 (1.1) | 0.16 (0.84) | 0.1 (0.6) |
| Triglyceride | 159 (126) | 145 (109) | 154 (122) | 152 (124) |
| Uric Acid | 6.0 (2.1) | 7.1 (2.7) | 6.6 (2.4) | 6.5 (2.4) |
| VLDL | 31 (16) | 28 (16) | 29 (16) | 28 (15) |
| eGFR | 58 (8) | 50 (15) | 54 (12) | 55 (12) |
| R Axis | 27 (27) | 18 (18) | 22 (50) | 22 (22) |
| PR Interval | 159 (159) | 175 (175) | 165 (212) | 165 (165) |
| P Axis | 47 (47) | 50 (50) | 48 (30) | 49 (49) |
| QRS Duration | 91 (91) | 110 (110) | 98 (25) | 96 (96) |
| QT | 392 (392) | 410 (410) | 400 (51) | 397 (397) |
| QTC | 434 (434) | 464 (464) | 445 (54) | 445 (445) |
| T Axis | 42 (42) | 70 (70) | 52 (53) | 51 (51) |
| Ventricular Rate | 76 (76) | 81 (81) | 77 (20) | 79 (79) |
| Avg RR Interval | 831 (831) | 794 (794) | 821 (194) | 809 (809) |
| Normal | 52.9% | 28.5% | 43.8% | 42.4% |
| Prior Infarct | 12.6% | 28.6% | 18.7% | 18.6% |
| Non-Spec T | 13.4% | 19.9% | 16.0% | 16.3% |
| Sinus Bradycardia | 15.4% | 10.7% | 14.1% | 13.1% |
| Non-Spec ST | 8.2% | 13.4% | 10.3% | 10.9% |
| Ischemia | 5.2% | 18.3% | 10.0% | 9.3% |
| Tachycardia | 7.9% | 8.0% | 7.5% | 9.2% |
| Left axis deviation | 5.9% | 14.8% | 9.3% | 9.2% |
| Atrial Fibrillation | 2.9% | 18.3% | 8.5% | 8.2% |
| LVH | 6.1% | 10.6% | 8.0% | 7.8% |
| Prior MI Ant. | 4.6% | 12.2% | 7.3% | 7.4% |
| PVC | 3.6% | 12.8% | 6.8% | 6.7% |
| First Deg Block | 3.9% | 9.3% | 6.3% | 6.4% |
| Right bundle branch block | 3.4% | 9.8% | 6.0% | 6.4% |
| Poor Tracing | 4.2% | 6.5% | 4.9% | 5.6% |
| PAC | 3.4% | 7.0% | 4.8% | 5.6% |
| Low QRS voltage | 3.5% | 6.2% | 4.4% | 5.1% |
| Prolonged QT | 3.2% | 8.4% | 5.0% | 4.9% |
| T-wave Inversion | 2.7% | 7.6% | 4.6% | 4.7% |
| Pacemaker | 1.2% | 10.0% | 4.6% | 4.0% |
| Incomplete RBBB | 3.2% | 3.0% | 3.1% | 3.5% |
| Fascicular Block | 2.0% | 4.9% | 3.2% | 3.4% |
| Left bundle branch block | 0.9% | 6.2% | 2.8% | 2.3% |
| Right axis deviation | 1.8% | 3.0% | 2.2% | 2.2% |
| Intraventricular Block | 0.8% | 5.2% | 2.3% | 1.9% |
| Atrial Flutter | 0.5% | 2.8% | 1.3% | 1.3% |
| Acute MI | 0.6% | 2.0% | 1.0% | 0.8% |
| Supraventricular Tachycardia | 0.3% | 0.8% | 0.4% | 0.5% |
| Early repolarization | 0.6% | 0.1% | 0.3% | 0.4% |
| incomplete left BBB | 0.1% | 0.9% | 0.4% | 0.4% |
| Second-degree AV block | 0.1% | 0.3% | 0.1% | 0.2% |
| Other Bradycardia | 0.1% | 0.2% | 0.1% | 0.1% |
| Complete Heart Block | 0.0% | 0.2% | 0.1% | 0.1% |
| Ventricular Tachycardia | 0.0% | 0.3% | 0.1% | 0.1% |

**Supplemental Table 8**: rECHOmmend model performance metrics (mean with 95% CI) across multiple threshold values. NPV, negative predictive value. PPV, positive predictive value.

| Threshold | NPV | PPV | Sensitivity | Specificity | Threshold |
| --- | --- | --- | --- | --- | --- |
| 0.1 | 93.8 [93.7,93.9] | 62.2 [61.3,63.0] | 72.7 [72.2,73.2] | 90.4 [90.1,90.6] | 0.1 |
| 0.2 | 91.7 [91.6,91.8] | 71.2 [70.3,72.0] | 60.5 [60.1,60.9] | 94.7 [94.5,94.8] | 0.2 |
| 0.3 | 90.2 [90.1,90.2] | 75.8 [75.1,76.5] | 51.8 [51.4,52.2] | 96.4 [96.3,96.5] | 0.3 |
| 0.4 | 89.0 [88.9,89.1] | 79.2 [78.3,80.0] | 44.7 [44.5,44.9] | 97.4 [97.3,97.5] | 0.4 |
| 0.5 | 88.0 [87.9,88.1] | 81.8 [81.2,82.4] | 38.6 [38.4,38.8] | 98.1 [98.1,98.2] | 0.5 |
| 0.6 | 87.1 [86.9,87.2] | 84.1 [83.3,84.8] | 32.8 [32.4,33.2] | 98.6 [98.6,98.7] | 0.6 |
| 0.7 | 86.1 [86.0,86.3] | 86.0 [85.3,86.7] | 26.9 [26.6,27.3] | 99.0 [99.0,99.1] | 0.7 |
| 0.8 | 85.0 [84.9,85.2] | 88.3 [87.5,89.1] | 19.7 [19.0,20.4] | 99.4 [99.4,99.5] | 0.8 |
| 0.9 | 83.3 [83.1,83.4] | 91.2 [90.5,91.9] | 8.0 [6.6,9.7] | 99.8 [99.8,99.9] | 0.9 |
| Youden | 95.6 [95.5,95.7] | 52.7 [52.3,53.2] | 82.2 [81.6,82.7] | 83.9 [83.5,84.4] | 0.05 |
| F1-score | 93.3 [93.1,93.4] | 64.8 [64.1,65.5] | 69.6 [68.8,70.5] | 91.8 [91.5,92.0] | 0.12 |
| F2-score | 96.3 [95.9,96.6] | 48.0 [46.0,50.0] | 85.9 [84.1,87.5] | 79.7 [77.6,81.8] | 0.04 |
| at 25% PPV | 99.1 [98.9,99.2] | 25.0 [25.0,25.0] | 98.4 [98.2,98.7] | 35.6 [34.8,36.4] | 0.01 |
| at 33% PPV | 98.2 [98.1,98.3] | 33.0 [33.0,33.0] | 95.2 [94.7,95.6] | 57.9 [57.3,58.5] | 0.02 |
| at 90% Spec | 94.0 [93.9,94.0] | 61.6 [61.1,62.0] | 73.5 [72.9,74.1] | 90.0 [90.0,90.0] | 0.1 |
| at 50% Sens | 89.9 [89.8,90.0] | 76.7 [75.8,77.6] | 50.0 [50.0,50.0] | 96.7 [96.6,96.8] | 0.32 |
| at 90% Sens | 97.1 [97.1,97.1] | 42.0 [41.4,42.6] | 90.0 [90.0,90.0] | 72.9 [72.4,73.4] | 0.03 |

**Supplemental Table 9**: Multi-site validation results. Performance of the rECHOmmend model using age, sex, and ECG traces as inputs, trained on data from Geisinger Medical Center only and tested at all other 11 clinical sites. AUROC, area under receiver operating curve. NPV, negative predictive value. PPV, positive predictive value.

|  | AUROC | NPV | PPV | Prevalence | Sensitivity | Specificity | Number of  patients |
| --- | --- | --- | --- | --- | --- | --- | --- |
| Lewistown | 0.898 | 98.5% | 24.6% | 10.6% | 91.60% | 66.70% | 39,805 |
| Holy Spirit | 0.912 | 98.7% | 27.3% | 10.8% | 92.10% | 70.50% | 53,014 |
| Jersey Shore Hospital | 0.912 | 98.4% | 32.2% | 14.7% | 93.60% | 66.10% | 2,263 |
| Shamokin Area  Community Hospital | 0.922 | 97.4% | 48.8% | 19.9% | 91.90% | 76.10% | 8,090 |
| Geisinger Commonwealth  School of Medicine | 0.820 | 87.6% | 54.4% | 38.9% | 88.20% | 52.90% | 6,884 |
| South Wilkes-Barre Hospital | 0.910 | 99.1% | 18.7% | 5.3% | 86.70% | 78.90% | 2,474 |
| Viewmont Imaging Center | 0.789 | 99.6% | 3.9% | 1.3% | 74.40% | 75.80% | 3,265 |
| Bloomsburg Hospital | 0.917 | 98.1% | 34.3% | 9.9% | 85.40% | 82.10% | 6,794 |
| Grays Woods Clinic | 0.856 | 95.1% | 38.7% | 14.8% | 76.30% | 79.10% | 28,043 |
| Scranton Community  Medical Center | 0.925 | 98.0% | 42.3% | 19.6% | 94.40% | 68.50% | 41,658 |
| Wyoming Valley  Medical Center | 0.915 | 98.0% | 36.6% | 14.3% | 90.90% | 73.90% | 123,573 |
| Aggregate (all other 11 sites combined) | 0.906 | 97.8% | 34.9% | 14.2% | 90.20% | 72.10% | 315,863 |

**Supplemental Table 10**: Cross-validation performance metrics (mean with 95% CI) computed with data prior to 2010. AUROC, area under receiver operating curve. AUPRC, area under precision-recall curve. PPV, positive predictive value.

|  | Prevalence | AUROC | AUPRC | PPV@90% Sens. | Spec.@90% Sens. |
| --- | --- | --- | --- | --- | --- |
| Aortic Stenosis | 1.3 [1.3,1.4] | 90.4 [89.4,91.4] | 12.8 [11.9,13.7] | 4.6 [3.9,5.4] | 74.7 [70.2,78.8] |
| Aortic Regurgitation | 1.4 [1.3,1.6] | 83.5 [82.6,84.3] | 8.4 [7.5,9.4] | 3.0 [2.5,3.5] | 56.6 [51.0,62.0] |
| Mitral Regurgitation | 3.6 [3.5,3.7] | 88.5 [87.9,89.1] | 25.4 [22.6,28.3] | 9.6 [8.8,10.4] | 68.1 [65.1,70.9] |
| Mitral Stenosis | 0.1 [0.1,0.2] | 89.5 [86.1,92.1] | 2.4 [1.7,3.3] | 0.4 [0.3,0.5] | 70.7 [59.0,80.2] |
| Tricuspid Regurgitation | 2.8 [2.7,2.9] | 89.9 [89.2,90.6] | 24.5 [21.5,27.6] | 8.6 [8.2,9.1] | 72.9 [71.6,74.2] |
| EF<50% | 7.0 [6.8,7.1] | 91.7 [91.1,92.2] | 48.5 [47.2,49.8] | 23.8 [22.4,25.3] | 78.4 [76.3,80.4] |
| IVS>15mm | 3.5 [3.4,3.6] | 82.9 [81.7,84.0] | 16.2 [15.3,17.1] | 7.0 [6.6,7.5] | 56.7 [53.2,60.1] |
| rECHOmmend | 13.3 [13.1,13.4] | 89.1 [88.8,89.5] | 57.4 [56.2,58.6] | 31.3 [29.9,32.8] | 69.8 [67.8,71.8] |

**Supplemental Table 11**: Severe disease only. Performance comparison of cross-validated models with varying input features for the composite endpoint (valve disease, reduced EF, increased IVS) using severe disease definitions only. All values are shown in percentage with the 95% CI in between brackets. Each model was tested on a random ECG per patient. AUROC, area under receiver operating curve. AUPRC, area under precision-recall curve. PPV, positive predictive value.

| Input Features | AUROC | PPV  @90% Sens. | AUPRC | Spec.  @90% Sens. |
| --- | --- | --- | --- | --- |
| A) Age + Sex | 78.9 [78.7,79.1] | 16.8 [16.4,17.1] | 31.6 [30.8,32.4] | 47.0 [46.6,47.5] |
| B) Demo, Labs, and Vitals | 86.1 [86.0,86.2] | 20.6 [20.3,21.0] | 53.4 [53.1,53.6] | 58.9 [58.5,59.3] |
| C) ECG Findings and Meas. | 89.5 [89.3,89.7] | 25.1 [24.8,25.5] | 58.9 [58.2,59.6] | 68.2 [67.7,68.7] |
| D) ECG Traces | 91.7 [91.5,91.8] | 30.3 [29.8,30.9] | 64.4 [63.7,65.1] | 75.5 [74.9,76.1] |
| Available from ECG system |  |  |  |  |
| Age + Sex + ECG Traces | 91.9 [91.7,92.1] | 31.2 [30.4,32.0] | 63.8 [62.9,64.6] | 76.4 [75.5,77.4] |
| C + D | 92.4 [92.2,92.6] | 32.4 [31.3,33.5] | 66.6 [66.1,67.0] | 77.7 [76.6,78.8] |
| Available from ECG + EHR |  |  |  |  |
| A + B + C | 92.3 [92.2,92.4] | 32.1 [31.4,32.8] | 67.5 [66.8,68.2] | 77.4 [76.9,78.0] |
| A + B + D | 93.3 [93.1,93.4] | 34.9 [34.3,35.6] | 70.5 [69.8,71.2] | 80.1 [79.5,80.7] |
| A + B + C + D | 93.6 [93.4,93.8] | 36.2 [35.2,37.1] | 71.2 [70.7,71.7] | 81.1 [80.3,81.9] |

**Supplemental Table 12**: Severe disease only. Age+Sex+ECG traces model results for cross-validation experiments for each individual outcome and rECHOmmend labels using severe disease definitions only. Results are shown at a random ECG per patient and averaged across 5 folds. All values are shown in percentage with the 95% CI in between brackets. AUROC, area under receiver operating curve. AUPRC, area under precision-recall curve. PPV, positive predictive value.

|  | Prevalence | AUROC | AUPRC | PPV  @90% Sens. | Spec.  @90% Sens. |
| --- | --- | --- | --- | --- | --- |
| Aortic Stenosis | 1.3 [1.3,1.4] | 92.6 [92.0,93.1] | 18.2 [17.2,19.4] | 5.9 [5.4,6.4] | 80.5 [78.3,82.5] |
| Aortic Regurgitation | 0.5 [0.5,0.5] | 83.6 [81.4,85.6] | 4.1 [4.0,4.3] | 0.9 [0.8,1.1] | 52.8 [44.4,61.0] |
| Mitral Regurgitation | 1.5 [1.5,1.6] | 91.4 [91.0,91.7] | 19.5 [18.3,20.8] | 5.8 [5.4,6.1] | 76.9 [76.0,77.7] |
| Mitral Stenosis | 0.1 [0.1,0.1] | 90.3 [87.1,92.8] | 1.9 [1.6,2.2] | 0.2 [0.1,0.4] | 68.9 [48.9,83.7] |
| Tricuspid Regurgitation | 1.3 [1.3,1.4] | 93.9 [93.5,94.2] | 27.0 [26.1,28.0] | 7.0 [6.5,7.6] | 84.0 [82.7,85.2] |
| EF<35% | 4.2 [4.1,4.2] | 95.9 [95.8,96.0] | 58.7 [57.2,60.3] | 27.9 [27.2,28.7] | 89.9 [89.6,90.3] |
| IVS>15mm | 4.0 [3.9,4.1] | 86.5 [86.2,86.8] | 22.1 [20.3,23.9] | 9.7 [9.5,9.9] | 65.1 [63.7,66.5] |
| rECHOmmend | 10.6 [10.5,10.7] | 91.9 [91.7,92.1] | 63.8 [62.9,64.6] | 31.2 [30.4,32.0] | 76.4 [75.5,77.4] |

**Supplemental Table 13**: Performance comparison of cross-validated models with varying input features for the composite endpoint (valve disease, reduced EF, increased IVS). All values are shown in percentage with the 95% CI in between brackets. Each model was tested on a random ECG per patient for TTE-confirmed only labels. AUROC, area under receiver operating curve. AUPRC, area under precision-recall curve. PPV, positive predictive value.

| Input Features | AUROC | PPV  @90% Sens. | AUPRC | Spec.  @90% Sens. |
| --- | --- | --- | --- | --- |
| A) Age + Sex | 75.7 [75.4,76.0] | 61.8 [61.3,62.3] | 78.0 [77.4,78.6] | 36.3 [35.7,36.9] |
| B) Demo, Labs, and Vitals | 81.4 [81.1,81.7] | 65.9 [65.3,66.6] | 83.8 [83.4,84.2] | 46.9 [45.9,47.9] |
| C) ECG Findings and Meas. | 85.1 [84.8,85.4] | 68.9 [68.2,69.5] | 87.1 [86.6,87.5] | 53.5 [52.7,54.4] |
| D) ECG Traces | 87.9 [87.8,88.1] | 73.5 [72.9,74.0] | 89.6 [89.2,89.9] | 62.9 [62.3,63.4] |
| Available from ECG System |  |  |  |  |
| Age + Sex + ECG Traces | 88.0 [87.7,88.2] | 74.0 [73.4,74.6] | 89.3 [89.0,89.6] | 63.8 [62.9,64.8] |
| C + D | 88.7 [88.5,88.9] | 74.6 [74.0,75.1] | 90.2 [89.8,90.5] | 64.9 [64.3,65.6] |
| Available from ECG + EHR |  |  |  |  |
| A + B + C | 88.0 [87.8,88.3] | 73.8 [73.3,74.3] | 89.5 [89.2,89.9] | 63.5 [63.0,64.0] |
| A + B + D | 88.8 [88.7,89.0] | 75.2 [74.7,75.6] | 90.3 [89.9,90.6] | 66.0 [65.6,66.5] |
| A + B + C + D | 89.3 [89.1,89.5] | 75.9 [75.3,76.5] | 90.7 [90.3,91.0] | 67.4 [66.7,68.0] |

**Supplemental Table 14**: Age+Sex+ECG traces model results for cross-validation experiments for each individual outcome and rECHOmmend. Results are shown at a random ECG per patient and averaged across 5 folds for TTE-confirmed only ECGs. All values are shown in percentage with the 95% CI in between brackets. AUROC, area under receiver operating curve. AUPRC, area under precision-recall curve. PPV, positive predictive value.

|  | Prevalence | AUROC | AUPRC | PPV  @90% Sens. | Spec.  @90% Sens. |
| --- | --- | --- | --- | --- | --- |
| Aortic Stenosis | 9.9 [9.5,10.2] | 86.7 [85.7,87.6] | 43.7 [42.1,45.3] | 21.8 [20.5,23.2] | 64.7 [61.6,67.7] |
| Aortic Regurgitation | 7.6 [7.3,7.8] | 78.8 [77.8,79.7] | 25.8 [24.3,27.4] | 11.8 [11.3,12.4] | 45.2 [43.3,47.1] |
| Mitral Regurgitation | 17.9 [17.4,18.4] | 86.7 [86.3,87.2] | 59.4 [58.1,60.8] | 35.2 [33.9,36.5] | 63.9 [62.9,64.8] |
| Mitral Stenosis | 1.1 [1.0,1.2] | 87.9 [86.1,89.5] | 9.5 [8.4,10.8] | 3.1 [2.7,3.6] | 68.6 [62.3,74.2] |
| Tricuspid Regurgitation | 18.7 [18.1,19.2] | 88.3 [87.4,89.0] | 66.1 [63.9,68.2] | 38.6 [36.6,40.6] | 67.1 [64.6,69.6] |
| EF<50% | 32.5 [32.4,32.6] | 90.2 [89.8,90.6] | 83.1 [82.3,83.9] | 60.3 [59.3,61.3] | 71.4 [70.2,72.6] |
| IVS>15mm | 16.1 [15.7,16.6] | 80.3 [79.7,81.0] | 43.8 [42.4,45.2] | 25.8 [24.7,27.0] | 50.3 [48.1,52.6] |
| rECHOmmend | 53.3 [53.0,53.7] | 88.0 [87.7,88.2] | 89.3 [89.0,89.6] | 74.0 [73.4,74.6] | 63.8 [62.9,64.8] |
